# Yoga Effect on Quality-of-Life Study Among Patients with Idiopathic Pulmonary Fibrosis (YES-IPF)

**DOI:** 10.1101/2025.05.20.25327762

**Authors:** Suha Kadura, Soumik Purkayastha, Josh Benditt, Amit Anand, Bridget Collins, Miguele De Quadros, Mafara Hobson, MJ Biswas, Lawrence Ho, Cathie Spino, Ganesh Raghu

**Author notes:** Corresponding Author: Ganesh Raghu, MD, University of Washington, Campus Box 356175, Seattle, WA 98195 USA.

## Abstract

**Rationale:** Patients with idiopathic pulmonary fibrosis have poor health-related quality of life.

**Objective:** Determine whether a modified yoga program in patients with idiopathic pulmonary fibrosis improves quality of life compared to usual care.

**Methods:** Randomized controlled, non-blinded, pilot clinical trial lasting 12 weeks with 2 arms involving 63 adults with idiopathic pulmonary fibrosis. The yoga program-arm consisted of interventions such as seated postures, breathing and meditation exercises designed by authors for idiopathic pulmonary fibrosis patients. The control arm continued with usual standard of care. Analysis of covariance was performed; no multiplicity adjustments were made on account of this being a pilot study.

**Measurements:** The primary outcomes were week-12 scores in seven patient-reported instruments, each with sub-domains, totaling 21 outcomes.

**Results:** 60 of the 63 participants (32 randomized to yoga; 31 receiving standard of care) completed the study (one death in each arm and one withdrawal in yoga group). Analysis of covariance for week-12 scores, adjusting for baseline scores and confounders, revealed significant treatment effects favoring yoga in the L-IPF cough domain (-9.29 points, 95% CI -18.37 to -0.20; p=0.045), in the L-IPF total score (-7.11, 95% CI -13.15 to -1.06; p=0.022), and in the R-scale-PF cough domain (-1.18, 95% CI - 2.27 to -0.10; p=0.034) in the study population.

**Conclusion:** Patients with idiopathic pulmonary fibrosis participating in a yoga program demonstrated significant improvement in quality of life assessed by the cough and total scores of L-IPF and the cough score of R-scale-PF than those receiving usual care.

**Summary:** We examine if the implementation of a modified yoga program improves patient-reported outcomes in patients with idiopathic pulmonary fibrosis. Our study reveals significant post-yoga improvements in the yoga group compared to the control group across different patient-reported outcomes. Such outcomes are meaningful endpoints in patients with idiopathic pulmonary fibrosis and serve as a critical endpoint in related clinical trials.

## 1. Introduction

Idiopathic pulmonary fibrosis carries significant mortality and symptom burden, with limited therapeutic options. Despite the discovery of antifibrotic agents that slow the rate of decline of forced vital capacity, patients living with idiopathic pulmonary fibrosis suffer from persistent respiratory symptoms and diminished quality of life. There are no available pharmacologic therapies for idiopathic pulmonary fibrosis that have been shown to impact patient-reported outcomes, such as symptoms or health-related quality-of-life. Though biomarkers such as forced vital capacity or image patterns may identify effects of interventions on biological pathways of the disease process, they likely do not fully encompass treatment impact on how patients ‘feel, function, and survive,’ and only weakly correlate with patient-reported symptom measures ^2–4^. Identification of treatment strategies for idiopathic pulmonary fibrosis with the aim of enhancing patients’ quality of life is a top priority.

Yoga is a mind-body practice that has become an increasingly popular and effective method to treat and manage symptoms of chronic diseases, and is the most commonly used complementary health approach in the United States ^5^. According to a 2017 national survey, about one in seven U.S. adults practiced yoga in the past 12 months, and the percentage of people practicing yoga grew from 2007 to 2017 ^6^. A mounting body of literature demonstrates the benefits of yoga not only in components of general wellness including stress, mental/emotional health, and sleep quality, but has also been shown to improve symptoms and quality of life in various chronic diseases including Chronic Obstructive Pulmonary Disease, asthma, cancer, heart failure, arthritis, and chronic pain ^7–12^.

Although classical yoga includes other elements, yoga as commonly practiced in the United States typically emphasizes standard components such as breathing exercises (pranayama), postures (asanas), and meditation (dhyana). Central to asanas and pranayama is diaphragmatic breathing, which may lead to improved chest expansion and decreased work of breathing. Studies evaluating the benefits of yoga in chronic respiratory diseases (largely asthma and Chronic Obstructive Pulmonary Disease) have shown improvement in variables such as forced expiratory volume in 1 second, tidal volume and minute ventilation, as well as increase in measures such as alveolar gas exchange and improved submaximal and maximal oxygen consumption, in addition to quality of life ^13–16^.

We sought to assess the effect of a standardized virtual, modified yoga program (henceforth, treatment) designed specifically for individuals with idiopathic pulmonary fibrosis at our center, with the effect on health-related quality of life as a primary objective. We hypothesized that a 12-week modified yoga program, designed specifically for individuals with idiopathic pulmonary fibrosis, would improve health- related quality of life compared to usual care.

## 2. Methods

### 2.1 Study design

This study was a non-blinded, single center, randomized pilot trial conducted by the Center for Interstitial Lung Diseases at the University of Washington Medical Center. It was approved by the Institutional Review Board at the University of Washington and registered with ClinicalTrials.gov (<u>NCT02848625</u>). See <u>Sections 2 to 6 of Online Data Supplement</u> for more details.

### 2.2 Participants

Consenting adults with idiopathic pulmonary fibrosis meeting criteria established in 2018 by the American Thoracic Society, European Respiratory Society, Japanese Respiratory Society, and Latin American Thoracic Society at time of screening were eligible ^17^. Exclusion criteria included comorbidities interfering with yoga (e.g., musculoskeletal discomfort), evaluation for lung transplant or hospice care, participation in yoga and/or pulmonary rehabilitation outside of the study at screening and during the study period. All patients were otherwise allowed to receive usual care for idiopathic pulmonary fibrosis including treatment with *nintedanib* or *pirfenidone* and supplemental oxygen when necessary, during the study period.

### 2.3 Randomization and masking

Eligible participants were randomly allocated in a 1:1 ratio to either the treatment arm (following the modified yoga program), or the control arm (receiving standard of care and following usual activities) with stratification by supplemental oxygen need prior to enrollment to account for disease severity. The randomization schedule was prepared using computer-generated permuted random blocks of size 4 for each stratum. The sequence of allocation was hidden from the investigators to prevent selection bias. Participants, investigators, and providers were not blinded to intervention; see <u>Figure E1 of the Online Data Supplement</u>.

### 2.4 Procedures

The modified yoga program consisted of seated postures, breathing, and meditation exercises designed by 5 certified yoga instructors tailored for fibrosis patients (see <u>Table E1 and Section 9 of Online Data Supplement</u>). Chair yoga, as used in our study, is a gentler yoga alternative for older adults. Classical yoga with more strenuous movement that may involve poses on the floor, significant joint and muscle stretching was considered not to be appropriate for the typical idiopathic pulmonary fibrosis patient population. Chair yoga is quite popular and many studios and training courses for its practice are available, with prior studies demonstrating benefit in elderly patients ^18^.

The protocol was intended to be in-person, though was amended due to the COVID-19 pandemic to reduce exposure risk, with participants in the treatment arm participating in group sessions conducted virtually via Zoom in individuals’ homes (5-6 per group). Sessions lasted one hour, twice/week, for 12 consecutive weeks, led by a certified yoga instructor. The same instructor led each group’s sessions. Participants received a secure link to a pre-recorded demo video to practice during their free time.

Participants in the treatment group received a journal at the beginning of the program and were encouraged to write in it at the end of each class (<u>see Section 8 of Online Data Supplement</u>). Journal entries were used in the qualitative assessment of the program. Attention control conditions were not implemented for the control group due to lack of feasibility and resource limitations during the pandemic. All patients were evaluated in person at baseline and after 12 weeks when lung function and 6-minute walk tests were performed, and patient-reported outcome measurements were collected.

### 2.5 Outcomes

The *primary outcomes* measured were total/domain-specific scores from seven validated health-related quality of life tools collected at baseline and at the end of the 12-week study period. These included the Living with Idiopathic Pulmonary Fibrosis symptoms module (L-IPF symptoms), a recently developed and validated visual analog scale (VAS) for IPF called Raghu scale for pulmonary fibrosis (R-Scale-PF), King’s Brief Interstitial Lung Disease questionnaire (K-BILD), EuroQol-Five-Dimensional Five-Level questionnaire (EQ-5D-5L), Hospital Anxiety and Depression (HAD) Scale, Patient-Reported Outcomes Measurement Information System Sleep Disturbance and Sleep-Related Impairment (PROMIS SD and SRI), and Epworth Sleepiness Scale (ESS) ^19–26^.

*Secondary outcomes* included changes from baseline to 12 weeks in absolute and percent predicted forced vital capacity (FVC), diffusing capacity of the lungs for carbon monoxide (DLCO), and 6-minute walk distance (6MWD).

### 2.6 Questionnaires for Patient Reported Outcomes

#### L-IPF questionnaire symptoms module

The L-IPF symptoms module contains 15 items, with the total score derived as the average of the dyspnea (7 items), cough (5 items), and energy/fatigue (3 items) domains, each with a 24-hour recall period ^19^. Each item is rated on a 5-point scale (0-4), with higher scores connoting greater impairment. The minimum clinically important difference estimates have been reported as 6 to 7 for the dyspnea domain score and 4 to 5 points for the cough domain score, though these estimates were investigated in a population of patients with progressive interstitial lung disease, other than idiopathic pulmonary fibrosis, which does not specifically apply to our study population ^27^.

#### R-scale-PF questionnaire

The R-scale-PF is a visual analog scale assessing 5 domains (cough, shortness of breath, fatigue, depressed mood, and overall sense of well-being). Each domain score ranges from 0 to 10 with 0.5 increments. The total score ranges from 0 to 50 with lower numbers indicating better health-related quality of life ^20^. A minimum clinically important difference estimate has not yet been defined in fibrosis patients.

Details for the remaining utilized questionnaires are provided in Section 7 of Online Data Supplement.

### 2.7 Statistical analysis

#### Study Design and Analysis Population

This pilot trial utilized patient-reported outcomes as primary outcomes, with sample size determined by feasibility rather than power calculations, as well-defined data on clinically important differences were not available for the fibrosis population in the implemented instruments. The primary analysis set was the modified intention-to-treat population defined as all randomized participants with a week 12 visit *Primary Analysis:* Between-group comparisons for outcome scores were conducted using analysis of covariance (ANCOVA), adjusting for baseline questionnaire scores, age, sex, BMI, and supplementary oxygen use. Any of the outcomes that demonstrated significant improvement in the study population was considered a favorable outcome. All analyses were conducted in R (version 4.4.1). Given this is a pilot trial, p-values were not adjusted for multiple comparisons. Missing data were handled via multiple imputation with chained equations. Sensitivity analyses using complete cases were performed.

## 3. Results

Between January 1-June 10, 2021, 63 consecutive, eligible patients provided consent. Thirty-two were randomly assigned to yoga and 31 to the control group. Thirty (94%) of 32 participants in the yoga group and 30 (97%) of 31 participants in the control group completed 12 weeks of assessment (Figure 1) to form the study population. One participant from each study arm was missing at least one week 12 questionnaires. DLCO data were not available for one participant at baseline and four at week 12, one did not have week 12 FVC data, and two participants were missing 6MWD at baseline and 4 at week 12.

**Figure 1:**
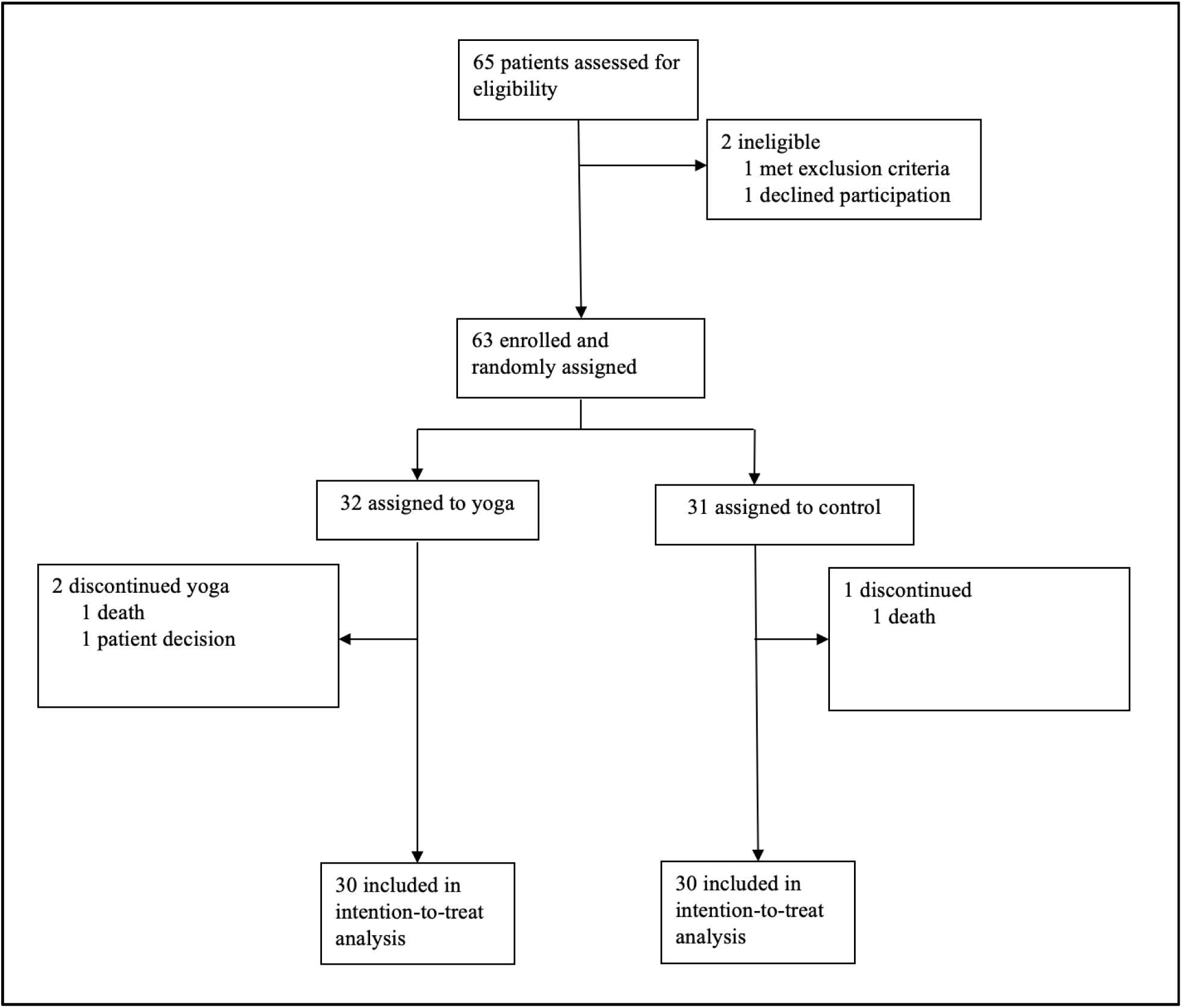
CONSORT diagram.

Baseline demographics are presented in Table 1. Most participants were male (41 [68%] of 60), with mean age of 73.4 [SD 8.21] years. Median disease duration was 3.2 years. Most characteristics are comparable between the treatment groups, with no clinically meaningful differences. Participants had mild to moderate impairment in lung function at baseline (mean FVC% predicted was 74.8 [SD 21.44] and % predicted DLCO was 54.0 [18.27]). Twenty-two (37%) participants used supplemental oxygen. Fifty-one (85%) participants were on background antifibrotic therapy with either *nintedanib* or *pirfenidone*. Comorbid conditions included obstructive sleep apnea in 28 (47%) participants, emphysema in 5 (8%), gastroesophageal reflux disease (confirmed by pH probe) in 25 (42%), and pulmonary hypertension (confirmed by right heart catheterization) in 11 (18%).

**Table 1.** Baseline characteristics of the study population.

|  | Control group<br>(N=30) | Treatment group<br>(N=30) | Overall<br>(N=60) |
| --- | --- | --- | --- |
| **Age (years)** |  |  |  |
| Mean (SD) | 74.3 (7.41) | 72.5 (8.97) | 73.4 (8.21) |
| **Body Mass Index (kg/m<sup>2</sup>)** |  |  |  |
| Mean (SD) | 26.5 (4.66) | 27.6 (4.55) | 27.1 (4.60) |
| **Sex, n (%)** |  |  |  |
| Female | 10 (33%) | 9 (30%) | 19 (32%) |
| Male | 20 (67%) | 21 (70%) | 41 (68%) |
| **Ethnicity, n (%)** |  |  |  |
| White | 27 (90%) | 27 (90%) | 54 (90%) |
| Other | 3 (10%) | 3 (10%) | 6 (10%) |
| **Current/former smoker? n (%)** |  |  |  |
| No | 19 (63%) | 16 (53%) | 35 (58%) |
| Yes | 11 (37%) | 14 (47%) | 25 (42%) |
| **Time since diagnosis (years)** |  |  |  |
| Median (Interquartile range) | 3.6 (2.93) | 3.0 (4.39) | 3.2 (3.67) |
| [25<sup>th</sup> percentile – 75<sup>th</sup> percentile] | [2.31 – 5.24] | [1.92 – 6.30] | [2.21 – 5.88] |
| **Surgical lung biopsy done? n (%)** |  |  |  |
| No | 17 (57%) | 19 (63%) | 36 (60%) |
| Yes | 13 (43%) | 11 (37%) | 24 (40%) |
| **FVC (L)** |  |  |  |
| Mean (SD) | 2.7 (0.77) | 2.9 (0.86) | 2.8 (0.82) |
| Median [25<sup>th</sup> percentile – 75<sup>th</sup> percentile] | 2.7 [2.02 – 2.96] | 3.1 [2.22 – 3.70] | 2.8 [2.08 – 3.37] |
| **FVC % predicted** |  |  |  |
| Mean (SD) | 70.0 (21.84) | 79.6 (20.28) | 74.8 (21.44) |
| Median [25<sup>th</sup> percentile – 75<sup>th</sup> percentile] | 70.5 [58.75 – 84.25] | 77.5 [66.75 – 94.75] | 73.0 [65.00 – 91.25] |
| **DLCO (mmol/min/kPa)** |  |  |  |
| Mean (SD) | 12.4 (3.41) | 13.3 (4.42) | 12.8 (3.93) |
| Missing | 0 (0%) | 1 (3.3%) | 1 (1.7%) |
| **DLCO % predicted value [\*]** |  |  |  |
| Mean (SD) | 52.5 (15.38) | 56.0 (20.97) | 54.0 (18.27) |
| Missing | 0 (0%) | 1 (3.3%) | 1 (1.7%) |
| **6MW distance (feet)** |  |  |  |
| Mean (SD) | 1311 (405.7) | 1269 (403.6) | 1290 (401.7) |
| Missing | 1 (3.3%) | 1 (3.3%) | 2 (3.3%) |
| **Supplemental oxygen use, n (%) [†]** |  |  |  |
| Supplemental O2 not required | 20 (67%) | 18 (60%) | 38 (63%) |
| Supplemental O2 required | 10 (33%) | 12 (40%) | 22 (37%) |
| **Obstructive sleep apnea reported, n (%)** |  |  |  |
| No | 19 (63%) | 13 (43%) | 32 (53%) |
| Yes, without PAP therapy | 4 (13%) | 10 (33%) | 14 (23%) |
| Yes, with PAP therapy | 7 (23%) | 7 (23%) | 14 (23%) |
| **Emphysema reported, n (%)** |  |  |  |
| No | 29 (97%) | 26 (87%) | 55 (92%) |
| Yes | 1 (3%) | 4 (13%) | 5 (8%) |
| **Antifibrotics, n (%)** |  |  |  |
| No | 3 (10%) | 6 (20%) | 9 (15%) |
| Yes | 27 (90%) | 24 (80%) | 51 (85%) |
| **Gastroesophageal reflux disease, n (%)** |  |  |  |
| No/Unknown | 13 (43%) | 15 (50%) | 28 (47%) |
| Yes, confirmed by pH probe | 14 (47%) | 11 (37%) | 25 (42%) |
| Yes, not confirmed by pH probe | 3 (10%) | 4 (13%) | 7 (12%) |
| **Pulmonary hypertension, n (%)** |  |  |  |
| No/Unknown | 22 (73%) | 16 (53%) | 38 (63%) |
| Yes, confirmed by right heart catheterization | 4 (13%) | 7 (23%) | 11 (18%) |
| Yes, not confirmed by right heart cath. | 4 (13%) | 7 (23%) | 11 (18%) |
| **Notes:**<br>[*] Percent predicted FVC and DLCO calculated using GLI reference equations<sup>47</sup>.<br>[†] Supplemental oxygen use is a stratification factor. |  |  |  |

In Table 2, adjusted comparisons between yoga and control groups for primary outcomes using multiple imputation are reported. For the L-IPF cough domain, the yoga group showed improvement from baseline score (mean 27.83 [SD = 25.69]) to week 12 (mean 18.10 [21.23]), while the control group remained stable (baseline: 24.83 [22.95]; week 12: 25.00 [22.09]). The imputation-based analysis showed a treatment effect that was statistically significant, favoring yoga (adjusted treatment effect: -9.29; 95% CI: [- 18.37, -0.20]; p=0.045). The yoga group showed improvement from baseline total L-IPF score (mean 30.84 [SD = 18.77]) to week 12 (mean 23.37 [16.49]), while the control group remained stable (baseline: 26.49 [19.60]; week 12: 27.16 [19.92]). The imputation-based analysis showed a treatment effect that was statistically significant, favoring yoga (adjusted treatment effect: -7.11; 95% CI: [-13.15, -1.06]; p=0.022). In the R-scale-PF cough domain, the yoga group demonstrated significant reduction in cough (baseline: 3.20 [SD 2.64]; week 12: 2.07 [2.31]), whereas the control group showed a slight worsening (baseline: 2.87 [2.21]; week 12: 3.14 [2.56]). The imputation-based analyses demonstrated significant treatment effects (-1.18; 95% CI: [-2.27, -0.10]; p=0.034).

**Table 2.**
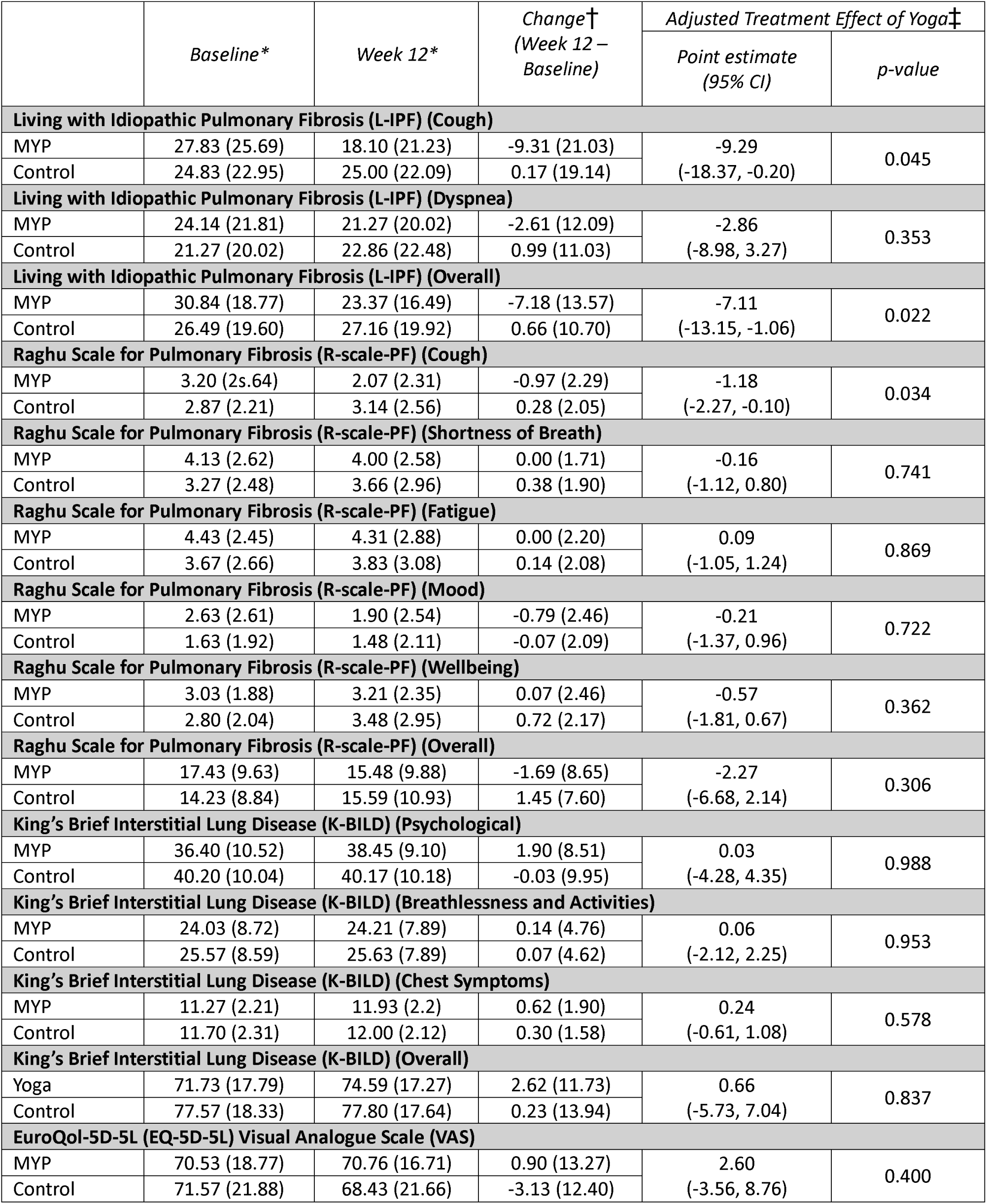
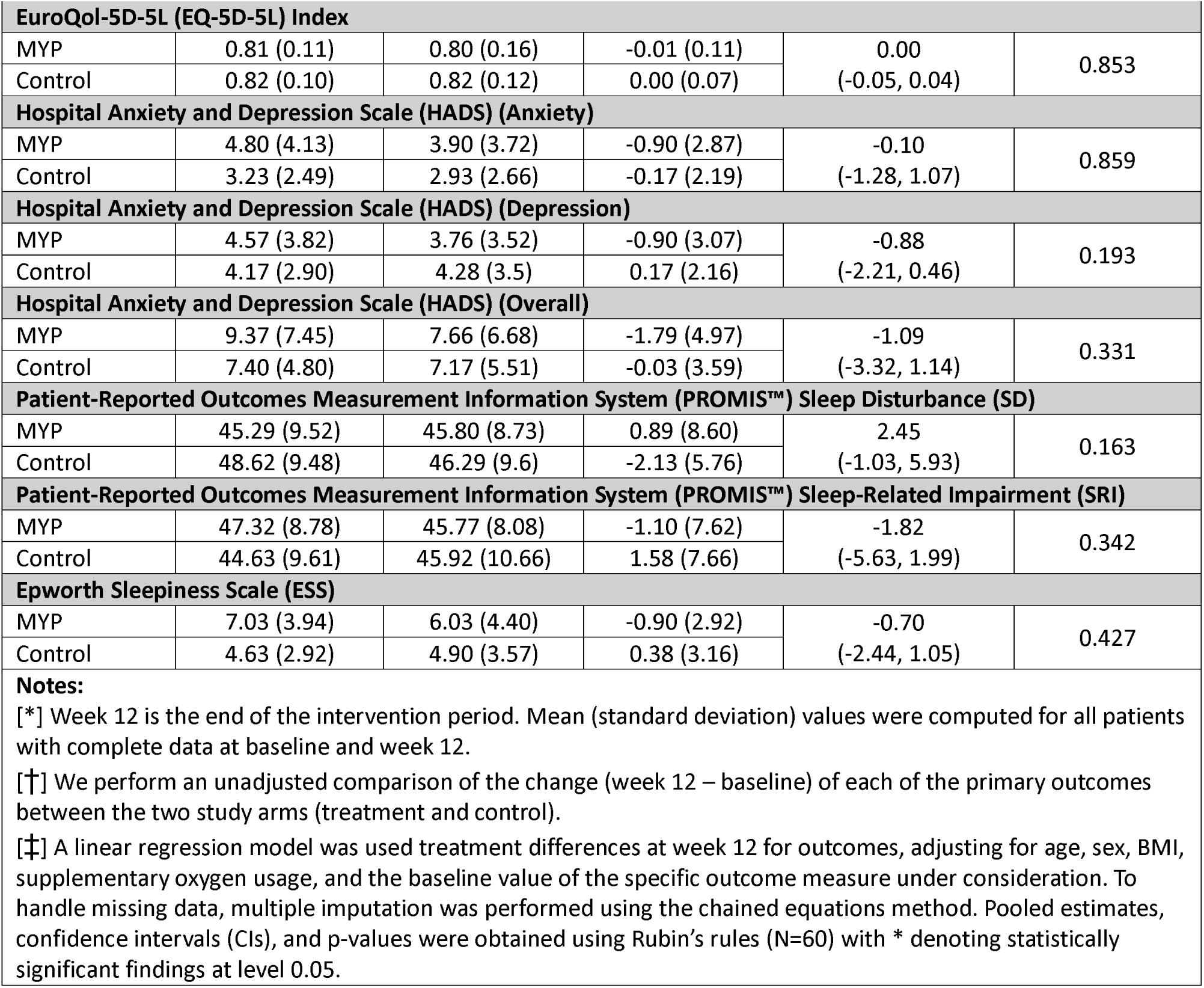
Changes from baseline to week 12 and adjusted treatment effect of modified yoga program in primary outcomes for the study population.

|  | Baseline* | Week 12* | Change†<br>(Week 12 –<br>Baseline) | Adjusted Treatment Effect of Yoga‡ |  |
| --- | --- | --- | --- | --- | --- |
|  |  |  |  | Point estimate<br>(95% CI) | p-value |
| Living with Idiopathic Pulmonary Fibrosis (L-IPF) (Cough) |  |  |  |  |  |
| MYP | 27.83 (25.69) | 18.10 (21.23) | -9.31 (21.03) | -9.29<br>(-18.37, -0.20) | 0.045 |
| Control | 24.83 (22.95) | 25.00 (22.09) | 0.17 (19.14) |  |  |
| Living with Idiopathic Pulmonary Fibrosis (L-IPF) (Dyspnea) |  |  |  |  |  |
| MYP | 24.14 (21.81) | 21.27 (20.02) | -2.61 (12.09) | -2.86<br>(-8.98, 3.27) | 0.353 |
| Control | 21.27 (20.02) | 22.86 (22.48) | 0.99 (11.03) |  |  |
| Living with Idiopathic Pulmonary Fibrosis (L-IPF) (Overall) |  |  |  |  |  |
| MYP | 30.84 (18.77) | 23.37 (16.49) | -7.18 (13.57) | -7.11<br>(-13.15, -1.06) | 0.022 |
| Control | 26.49 (19.60) | 27.16 (19.92) | 0.66 (10.70) |  |  |
| Raghu Scale for Pulmonary Fibrosis (R-scale-PF) (Cough) |  |  |  |  |  |
| MYP | 3.20 (2s.64) | 2.07 (2.31) | -0.97 (2.29) | -1.18<br>(-2.27, -0.10) | 0.034 |
| Control | 2.87 (2.21) | 3.14 (2.56) | 0.28 (2.05) |  |  |
| Raghu Scale for Pulmonary Fibrosis (R-scale-PF) (Shortness of Breath) |  |  |  |  |  |
| MYP | 4.13 (2.62) | 4.00 (2.58) | 0.00 (1.71) | -0.16<br>(-1.12, 0.80) | 0.741 |
| Control | 3.27 (2.48) | 3.66 (2.96) | 0.38 (1.90) |  |  |
| Raghu Scale for Pulmonary Fibrosis (R-scale-PF) (Fatigue) |  |  |  |  |  |
| MYP | 4.43 (2.45) | 4.31 (2.88) | 0.00 (2.20) | 0.09<br>(-1.05, 1.24) | 0.869 |
| Control | 3.67 (2.66) | 3.83 (3.08) | 0.14 (2.08) |  |  |
| Raghu Scale for Pulmonary Fibrosis (R-scale-PF) (Mood) |  |  |  |  |  |
| MYP | 2.63 (2.61) | 1.90 (2.54) | -0.79 (2.46) | -0.21<br>(-1.37, 0.96) | 0.722 |
| Control | 1.63 (1.92) | 1.48 (2.11) | -0.07 (2.09) |  |  |
| Raghu Scale for Pulmonary Fibrosis (R-scale-PF) (Wellbeing) |  |  |  |  |  |
| MYP | 3.03 (1.88) | 3.21 (2.35) | 0.07 (2.46) | -0.57<br>(-1.81, 0.67) | 0.362 |
| Control | 2.80 (2.04) | 3.48 (2.95) | 0.72 (2.17) |  |  |
| Raghu Scale for Pulmonary Fibrosis (R-scale-PF) (Overall) |  |  |  |  |  |
| MYP | 17.43 (9.63) | 15.48 (9.88) | -1.69 (8.65) | -2.27<br>(-6.68, 2.14) | 0.306 |
| Control | 14.23 (8.84) | 15.59 (10.93) | 1.45 (7.60) |  |  |
| King’s Brief Interstitial Lung Disease (K-BILD) (Psychological) |  |  |  |  |  |
| MYP | 36.40 (10.52) | 38.45 (9.10) | 1.90 (8.51) | 0.03<br>(-4.28, 4.35) | 0.988 |
| Control | 40.20 (10.04) | 40.17 (10.18) | -0.03 (9.95) |  |  |
| King’s Brief Interstitial Lung Disease (K-BILD) (Breathlessness and Activities) |  |  |  |  |  |
| MYP | 24.03 (8.72) | 24.21 (7.89) | 0.14 (4.76) | 0.06<br>(-2.12, 2.25) | 0.953 |
| Control | 25.57 (8.59) | 25.63 (7.89) | 0.07 (4.62) |  |  |
| King’s Brief Interstitial Lung Disease (K-BILD) (Chest Symptoms) |  |  |  |  |  |
| MYP | 11.27 (2.21) | 11.93 (2.2) | 0.62 (1.90) | 0.24<br>(-0.61, 1.08) | 0.578 |
| Control | 11.70 (2.31) | 12.00 (2.12) | 0.30 (1.58) |  |  |
| King’s Brief Interstitial Lung Disease (K-BILD) (Overall) |  |  |  |  |  |
| Yoga | 71.73 (17.79) | 74.59 (17.27) | 2.62 (11.73) | 0.66<br>(-5.73, 7.04) | 0.837 |
| Control | 77.57 (18.33) | 77.80 (17.64) | 0.23 (13.94) |  |  |
| EuroQol-5D-5L (EQ-5D-5L) Visual Analogue Scale (VAS) |  |  |  |  |  |
| MYP | 70.53 (18.77) | 70.76 (16.71) | 0.90 (13.27) | 2.60<br>(-3.56, 8.76) | 0.400 |
| Control | 71.57 (21.88) | 68.43 (21.66) | -3.13 (12.40) |  |  |

| EuroQol-5D-5L (EQ-5D-5L) Index |  |  |  |  |  |
| --- | --- | --- | --- | --- | --- |
| MYP | 0.81 (0.11) | 0.80 (0.16) | -0.01 (0.11) | 0.00<br>(-0.05, 0.04) | 0.853 |
| Control | 0.82 (0.10) | 0.82 (0.12) | 0.00 (0.07) |  |  |
| Hospital Anxiety and Depression Scale (HADS) (Anxiety) |  |  |  |  |  |
| MYP | 4.80 (4.13) | 3.90 (3.72) | -0.90 (2.87) | -0.10<br>(-1.28, 1.07) | 0.859 |
| Control | 3.23 (2.49) | 2.93 (2.66) | -0.17 (2.19) |  |  |
| Hospital Anxiety and Depression Scale (HADS) (Depression) |  |  |  |  |  |
| MYP | 4.57 (3.82) | 3.76 (3.52) | -0.90 (3.07) | -0.88<br>(-2.21, 0.46) | 0.193 |
| Control | 4.17 (2.90) | 4.28 (3.5) | 0.17 (2.16) |  |  |
| Hospital Anxiety and Depression Scale (HADS) (Overall) |  |  |  |  |  |
| MYP | 9.37 (7.45) | 7.66 (6.68) | -1.79 (4.97) | -1.09<br>(-3.32, 1.14) | 0.331 |
| Control | 7.40 (4.80) | 7.17 (5.51) | -0.03 (3.59) |  |  |
| Patient-Reported Outcomes Measurement Information System (PROMIS™) Sleep Disturbance (SD) |  |  |  |  |  |
| MYP | 45.29 (9.52) | 45.80 (8.73) | 0.89 (8.60) | 2.45<br>(-1.03, 5.93) | 0.163 |
| Control | 48.62 (9.48) | 46.29 (9.6) | -2.13 (5.76) |  |  |
| Patient-Reported Outcomes Measurement Information System (PROMIS™) Sleep-Related Impairment (SRI) |  |  |  |  |  |
| MYP | 47.32 (8.78) | 45.77 (8.08) | -1.10 (7.62) | -1.82<br>(-5.63, 1.99) | 0.342 |
| Control | 44.63 (9.61) | 45.92 (10.66) | 1.58 (7.66) |  |  |
| Epworth Sleepiness Scale (ESS) |  |  |  |  |  |
| MYP | 7.03 (3.94) | 6.03 (4.40) | -0.90 (2.92) | -0.70<br>(-2.44, 1.05) | 0.427 |
| Control | 4.63 (2.92) | 4.90 (3.57) | 0.38 (3.16) |  |  |
| **Notes:** |  |  |  |  |  |
| [*] Week 12 is the end of the intervention period. Mean (standard deviation) values were computed for all patients with complete data at baseline and week 12. |  |  |  |  |  |
| [†] We perform an unadjusted comparison of the change (week 12 – baseline) of each of the primary outcomes between the two study arms (treatment and control). |  |  |  |  |  |
| [‡] A linear regression model was used treatment differences at week 12 for outcomes, adjusting for age, sex, BMI, supplementary oxygen usage, and the baseline value of the specific outcome measure under consideration. To handle missing data, multiple imputation was performed using the chained equations method. Pooled estimates, confidence intervals (CIs), and p-values were obtained using Rubin’s rules (N=60) with * denoting statistically significant findings at level 0.05. |  |  |  |  |  |

Figure 2 illustrates individual patient trajectories from baseline to week 12 across three pulmonary fibrosis outcome measures through a waterfall plot.

**Figure 2:**
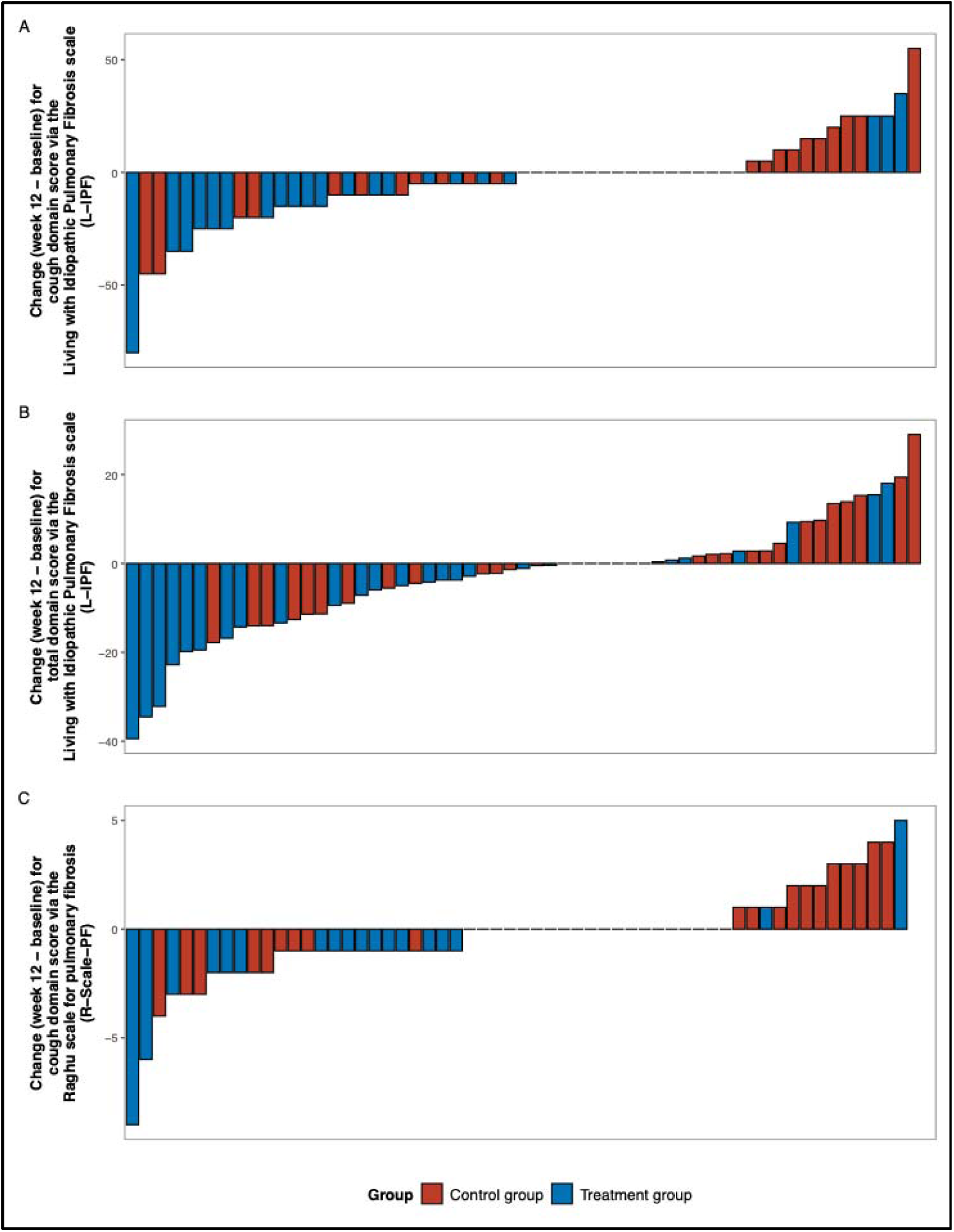
Waterfall plot to illustrate individual patient changes from baseline to week 12 across three pulmonary fibrosis outcome measures. Panel (A) shows changes in the cough domain score using the Living with Idiopathic Pulmonary Fibrosis (L-IPF) scale; (B) presents changes in the total domain score via the L- IPF scale, and (C) displays changes in the cough domain score using the Raghu Scale for Pulmonary Fibrosis (R-Scale-PF).

In <u>Table E2 of Online Data Supplement</u>, unadjusted and adjusted comparison between the yoga and control groups for secondary outcomes are shown using the multiple imputation approach. There were no significant differences in any of the secondary outcomes over 12 weeks. Findings from the sensitivity analyses are presented in <u>Table E3 of Online Data Supplement</u>.

A qualitative content analysis was performed through participants’ journals. Central themes identified leading to sense of improvement in overall well-being included social connection, managing fibrosis symptoms, changed mindset (<u>Figure E2 of Online Data Supplement</u>). Regarding social connection, participants reported feeling a sense of community and support during the program. Participants felt less self-conscious about experiencing symptoms (such as cough) and enjoyed speaking with others about their experiences and being able to monitor their progress alongside others who shared their diagnosis. Several participants reported incorporating more deliberate and controlled breathing techniques into daily activities, with improved knowledge and understanding of the connection between mind and body. From a mindset standpoint, participants reported feeling an increase in confidence in their ability and motivation to increase physical activity and push themselves despite their debilitating diagnosis.

## 4. Discussion

This pilot trial provides evidence that participants receiving 12-week modified yoga program experienced improvements in health-related quality-of-life outcomes, particularly in the cough domain scores of both the L-IPF and R-Scale-PF questionnaires, compared to those who continued usual daily activities. The strength of this conclusion is bolstered by consistency of results showing greater improvements in multiple quality of life instruments—including HADS, K-BILD, total R-scale-PF score, PROMIS SRI and ESS, following intervention in the yoga group than the control group.

To our knowledge, this is the first idiopathic pulmonary fibrosis clinical trial reporting improvement in outcomes in the intervention group as primary endpoints. As emphasized by a recent landmark international symposium involving patients, academic investigators, and regulatory representatives, endpoints in idiopathic pulmonary fibrosis clinical trials should resonate more closely with the tangible patient experience ^2^. The integration of adequately validated outcomes is an important step in this direction. The statistically significant improvement in the L-IPF cough domain score (and numeric improvement in KBILD score) in the yoga arm participants in our study is clinically meaningful to patients, as both of these questionnaires were developed with input from idiopathic pulmonary fibrosis patients and have undergone extensive validity and reliability testing ^2,21,28^. In a patient population with progressive interstitial lung disease, other than idiopathic pulmonary fibrosis, Swigris et al. provided estimates of 4 to 5 points in the L-PF cough domain score as a clinically meaningful threshold ^27^. Other scale tools, such as cough visual analogue scale, carry prognostic and quality of life value in fibrosis and are being increasingly used in interventional trials in idiopathic pulmonary fibrosis ^32,33^.

Previous studies have demonstrated a clear benefit of yoga in other chronic respiratory diseases on validated health-related quality-of-life scores, including asthma and COPD ^8,9,34,35^. Our study is the first to evaluate the effect of yoga on patients with idiopathic pulmonary fibrosis with positive findings despite the barrier of virtual rather than in-person sessions due to the COVID-19 pandemic, particularly since previous data suggests participant preference for in-person yoga classes ^36^. Two unpublished results showed mixed results regarding improvements in quality of life scores following participation in yoga, although both were designed as feasibility/safety studies with less than 20 participants each (one study in idiopathic pulmonary fibrosis patients and another in patients with fibrotic ILD of any cause) ^37,38^. We did not find statistically significant improvement in anxiety and depression scores measured by the HADS in contrast to a previous study that evaluated a general group of chronic lung disease patients awaiting lung transplant ^39^.

Although numerically greater improvements in the ESS and PROMIS SRI scores were observed in the yoga group, our study did not show statistically significant benefit in sleep quality despite prior reports showing improvement in sleep quality and insomnia with yoga participation in other patient populations ^40^. There is a complicated relationship with sleep quality in idiopathic pulmonary fibrosis patients, in whom comorbid sleep related breathing disorders such as OSA and insomnia are common and likely worsen the quality of life and may even increase mortality in patients with idiopathic pulmonary fibrosis ^41–43^. Another factor reported to be potentially contributing to increased arousal index in idiopathic pulmonary fibrosis is comorbid gastroesophageal reflux disease ^41^. Further prospective studies are needed to better evaluate the effects of yoga on overall sleep quality in idiopathic pulmonary fibrosis patients.

Possibilities of the potential mechanism for improved quality of life in fibrosis patients participating in yoga include improved posture, allowing more efficient use of abdominal and diaphragmatic muscles. Various yoga exercises such as deep diaphragmatic breathing can improve vagal tone, which may help diminish enhanced cough reflex sensitivity as previously described in patients with idiopathic pulmonary fibrosis ^44^.

The results of this study must be interpreted in the context of several limitations amidst its strengths. First, this was a single center, pilot trial designed to assess feasibility with the aim of informing future studies with adequate sample size calculations and powered to detect significant differences in trial outcomes between groups. This was an exploratory pilot trial designed primarily to assess feasibility and generate hypotheses for future studies. We acknowledge that with multiple endpoints (21 in total) and no adjustment for multiple comparisons, our findings are vulnerable to Type I error. However, the consistency of findings across multiple cough-related measures strengthens our confidence in these specific results. Future confirmatory trials should pre-specify primary endpoints and include appropriate statistical adjustments. A second limitation is the relatively small sample size and study duration of only 12 weeks. Third, since a blinded trial is not possible, it is difficult to distinguish between yoga-specific and nonspecific benefits (e.g. expectation bias, social connection during class, interpersonal skills of the yoga instructor). Finally, future trials should include well-validated measures of cough such as The Leicester Cough Questionnaire ^45,46^.

In summary, this is the first idiopathic pulmonary fibrosis clinical trial demonstrating improvement in patient-reported outcomes as primary endpoints, and the first study to evaluate the effect of yoga practice in this patient population. Besides demonstrating safety and feasibility of a yoga program that patients can engage in from the comfort of their homes, the study demonstrated improved health-related quality of life measures in patients with idiopathic pulmonary fibrosis following participation in a yoga program supervised by instructors. These results are clinically significant given the critical need for therapeutic options that improve outcomes that are meaningful to patients suffering with this disease. A larger, multi-center trial is warranted to further establish improved health-related quality of life via participation in a yoga program tailored for patients with idiopathic pulmonary fibrosis, in keeping with an emphasis on patient-centered outcomes.

## Contributions

1. *Suha Kadura:* Data gathering, interpretation of result, writing first draft, revised versions and collated inputs from all authors of the manuscript.
2. *Soumik Purkayastha:* Data analyses, interpretation of result, writing first draft, revised versions and collated inputs from all authors of the manuscript.
3. *Josh Benditt and Amit Anand:* Developed the protocol and study design for the modified yoga program /exercises used in the trial; reviewed the data analyzed, interpreted results.
4. *Josh Benditt, Amit Anand, Miguele De Quadros, Mafara Hobson, and MJ Biswas:* Certified yoga instructors for patients.
5. *Bridget Collins:* Enrolled consenting participants in the study, participated in study design, interpretation of results, reviewing manuscript and provided input.
6. *Lawrence Ho:* Enrolled and encouraged patients to participate in the trial.
7. *Cathie Spino:* Reviewed data analyses and interpretation, reviewed manuscript and provided input.
8. *Ganesh Raghu:* designed the trial and study protocol, enrolled consenting participants in the study, interpretation of results, supervised the conduct of trial and execution of the trial; manuscript writing.

All authors reviewed the drafts of manuscript, provided input, contributed to writing the manuscript and approved the final manuscript for submission.

## Data availability statement

In accordance with our protocol registered at ClinicalTrials.gov (NCT02848625), individual participant data from this study will not be made available publicly. This approach was established to protect participant privacy and confidentiality as stated in our informed consent process. Summary data supporting the findings of this study are available within the article and its supplementary materials. Researchers interested in collaborative analyses may contact the corresponding author with specific research questions and proposed methodologies for consideration.

