## Supplementary material for "Yoga Effect on Quality-of-Life Study Among Patients with Idiopathic Pulmonary Fibrosis (YES-IPF)": Online Data Supplement

1. **Study title**

Yoga effect on quality-of-life among patients with idiopathic pulmonary fibrosis (YES-IPF): A prospective, randomized control pilot trial by virtual means during the COVID-19 pandemic in Seattle, USA. ClinicalTrials.gov registration: NCT02848625 [[**link**](https://clinicaltrials.gov/study/NCT02848625?cond=ipf&intr=yoga&rank=1)]

2. **Background and Rationale**

There has been ongoing debate regarding clinically meaningful endpoints for clinical trials involving patients with Idiopathic Pulmonary Fibrosis (IPF). Measurements of pulmonary function (forced vital capacity or FVC and diffusion capacity for Carbon Monoxide or DLCO) are often used, although it is often argued that assessing for change in quality of life measures and/or mortality would be more meaningful, patient centered outcomes ^1^.

There is only one study to date regarding the effect of yoga on the quality of life and physiologic measures of pulmonary function among patients with IPF. Fell et al presented an abstract in 2011 where patients with IPF participated in biweekly 75-minute yoga sessions for 4 weeks. At the end of 4 weeks, there were no significant changes in health-related quality of life (HRQoL) as measured by the SGRQ and SF-36 or in 6MWD. However, this study did demonstrate that a yoga program may be safe and feasible among patients with IPF. Yoga has been studied in other chronic lung diseases. Studies of the effect of yoga and associated breathing techniques on symptoms and physiologic parameters in patients with asthma have drawn variable conclusions ^2,3^. Santana et al studied the effects of Iyenger yoga practice on patients with chronic lung disease awaiting lung transplant, 20% of whom had IPF, and found that anxiety (by the Hospital Anxiety and Depression Score) and fatigue (Chronic Respiratory Questionnaire) scores were improved after 12 weeks ^4^. A meta-analysis of 5 randomized controlled trials of COPD patients participating in yoga training demonstrated some improvement in FEV1 and 6MWD, although quality of life measures were not assessed ^5^. This meta-analysis does, however, suggest that yoga programs can be completed safely among patients with chronic lung disease. While participation in pulmonary rehabilitation programs has been shown to improve dyspnea and quality of life in patients with IPF, and is in fact recommended in evidence based guidelines, the effect of complementary therapies requiring focused breathing techniques, such as yoga, on quality of life, dyspnea and pulmonary function in this patient population has not yet been studied ^6–9^.

Previous studies measuring quality of life in patients with IPF have used the St George’s Respiratory Questionnaire (SGRQ), the short form 36 health survey (SF-36), EuroQol instrument, and A Tool to Assess Quality of life in IPF (ATAQ-IPF) ^10^. In terms of differences within the population of IPF patients, the initial study for ATAQ-IPF found that scores were significantly different among patients who required supplemental O2 and those who did not, with greater impairment among the former ^11^.

We propose to perform a pilot study of the effect of yoga on quality of life among patients with IPF when compared to usual care. While Fell et al did not observe a significant change in quality of life, their program was only 4 weeks long, and there was not a control group. We will also measure any effect of yoga on pulmonary function measured by PFTs and 6MWD.

3. **Objectives**

*Primary:* Identify whether routine a modified yoga program/exercises 2 times per week x 12 weeks is associated with improved quality of life (measured by any of the 7 validated tools to assess HRQOL) compared to usual care among patients with IPF. Questionnaires that will be used include:

1. Living with Idiopathic Pulmonary Fibrosis questionnaire symptoms module (L-IPF symptoms).
2. Raghu scale for pulmonary fibrosis, or R-Scale-PF.
3. Hospital Anxiety and Depression (HAD) Scale.
4. King’s Brief Interstitial Lung Disease questionnaire (K-BILD).
5. EuroQol-Five-Dimensional Five-Level questionnaire (EQ-5D-5L).
6. Epworth Sleepiness Scale (ESS).
7. Patient-Reported Outcomes Measurement Information System Sleep Disturbance and Sleep-Related Impairment (PROMIS SD and SRI).

*Secondary:* Determine whether IPF patients participating in MYP via Zoom classes x 12 weeks have

1. Improvement in FVC or less decline in FVC compared to control patients (usual care).
2. Improvement in 6-minute walk distance or less decline in 6-minute walk distance compared to control patients (usual care)
4. **Trial Design:** Randomized controlled trial (nonblinded)

*Study population:* Patients with IPF with diagnosis in accordance with 2018 ATS/ERS/JRS/ALAT criteria for diagnosis and management of IPF seen at the Center for ILD, UWMC.

*Outcomes*

1. Primary outcome: Change in QOL as measured by any of the 7 HRQOL questionnaires.
2. Secondary outcomes: % change in FVC over 12 weeks, % change in 6-minute walk distance (vs actual change in distance) over 12 weeks.

*Exposure:* Yoga 2 times per week, 1 hour per session, for 12 weeks through the Modified Yoga Program. The Modified Yoga Program consists of:

1. Pranayama (breathing exercises)
2. Asanas (seated yoga postures)
3. Dhyana (guided meditation)
4. Taught by Certified Yoga Teacher (CYT)
5. 2 x/week for 12 weeks

Participants received a secure [link](https://vimeo.com/514801390) to a pre-recorded demo video to practice during their free time.

*Study population:*

1. Include: Consecutive consented patients with IPF diagnosis seen in UWMC CILD
2. Exclude:
   a. Patients with active malignancy
   b. Comorbidities that would prohibitive: paralysis, musculoskeletal discomfort or broken limbs, etc.
   c. Patients being evaluate for lung transplant or receiving hospice care
   d. Patients regularly participating in yoga and/or pulmonary rehabilitation outside of the study at screening and during the study period
5. **Methodology:**

*Sample size:*

1. Acknowledging that this is the first study utilizing PROs as a primary outcome measure and was undertaken as a pilot trial, the study sample size was chosen based on feasibility considerations without formal power calculations.
2. Plan to randomize 60 eligible participants
3. Could consider power calculation based on KBILD MCID (MCID for KBILD in IPF is 3.9 points). Remaining PROs do not have clearly defined MCID for IPF.

*Randomization method:*

Eligible participants to be randomly allocated in a 1:1 ratio to yoga program or continuing “usual day-to-day activities,” with stratification by need to have supplemental oxygen prior to study enrollment to account for disease severity.

1. Study statistician to prepare randomization schedule using computer-generated permuted random blocks of size 4 for each stratum.
2. Sequence of allocation hidden from investigators to prevent selection bias.

*Potential confounding factors*: Sex, Age, Prior or current experience with yoga, Weight, and Concomitant lung disease (mainly COPD or asthma)

*Patient recruitment and consent:*

1. Recruitment by nurse coordinators or study investigators.
2. Consent by study RNs
6. **Statistical analysis plan:**

*Sample size/power calculation:* The sample size for this pilot trial was determined based on feasibility, as MCIDs for HRQoL instruments in IPF are not well established. Given the exploratory nature of the study, hypothesis testing was conducted without p-value adjustment for multiple endpoints. Point estimates with 95% confidence intervals (CIs) were reported to compare treatment effects. The primary analysis set was the modified intention-to-treat population, defined as all randomized participants with a week 12 visit.

*Exploratory analysis and modeling:* Descriptive statistics were presented for baseline, week 12, and change-from-baseline values in primary and secondary outcomes. For continuous variables, means with standard deviations (SDs) or medians with interquartile ranges were reported; categorical variables were summarized as frequencies and percentages. Between-group comparisons for week 12 outcome scores were conducted using ANCOVA, adjusting for baseline outcome scores, age, sex, BMI, and supplementary oxygen use. All statistical analyses were performed in R, version 4.4.1.

*Missing Data Handling:* Missing data were addressed using multiple imputation with chained equations (MICE) using the mice package version 3.17.0 ^12^. One hundred imputations were generated using Classification and Regression Trees (CART) as the imputation method. The imputation model included sex, BMI, age, and treatment arm as predictors, selected to ensure that all variables relevant to the analysis model were incorporated in the imputation process. Each imputed dataset was analyzed separately using the pre-specified linear regression model with post-treatment score as the outcome, adjusted for baseline measurements, sex, BMI, age, oxygen levels, and treatment arm. Results from all 100 analyses were then pooled according to Rubin's rules to account for both within- and between-imputation variability ^13^. This approach provides unbiased parameter estimates and correct standard errors under the missing at random (MAR) assumption. Stability of the results was supported by the large number of imputations utilized, which exceeds standard recommendations for complex analyses ^14,15^.

*Sensitivity Analysis:* Sensitivity analyses using complete cases were performed, applying the same ANCOVA model but including only participants with complete data for all variables. The robustness of the ANCOVA approach was assessed by checking the assumptions of normality of residuals (Shapiro-Wilk test), homogeneity of variance (Bartlett's test), and homogeneity of regression slopes (testing the interaction between baseline measurement and treatment arm). While 6 of the 21 outcomes examined demonstrated slight deviations from normality, our balanced design with adequate sample size (n=30 per group) ensures the robustness of ANCOVA to mild violations of normality ^16^. The other ANCOVA assumptions were adequately met.

7. **Description of questionnaires for PROs not included in main text:**

*K-BILD questionnaire:* K-BILD contains 15 items, a seven-point Likert scale and three domains (breathlessness and activities, psychological, and chest symptoms). The K-BILD domain and total score ranges are 0–100; 100 represents best health status. The MCID for the total K-BILD score is estimated to be 3.9 points in patients with ILD and IPF ^17^ .

*EQ-5D-5L:* The EQ-5D-5L assesses five dimensions including mobility, self-care, usual activities, pain/discomfort and anxiety/depression. Each domain has five possible responses that are combined to form a sum utility score which is then converted into a specific index value using a country-specific set (in our case USA in English). Those completing the questionnaire also rate their overall health on a 20 cm VAS with a score of 0 representing “the worse health you can imagine” and 100 representing “the best health you can imagine”^18^. The MCID estimate for the EQ-5D-5L in patients with fibrotic ILD, including IPF, ranges from 0.0050 to 0.054 and from 0.078 to 0.095 for the anchor-based and distribution-based methods, respectively ^19^.

*HADS:* The HADS is a self-reporting rating scale of 14 items on a 4-point Likert scale (range 0-3). It is designed to measure anxiety and depression (7 items for each domain). The total score is the sum of the 14 items, and for each domain the score is sum of the respective 7 items (ranging from 0-21). A higher score indicates higher distress ^20^. There is not an established MCID for the HADS questionnaire in IPF patients.

*PROMIS SD and SRI:* Sleep disturbance and sleep-related impairment are measured using PROMIS 8-item short forms. Both measures use a 5-point Likert scale. Each raw score is converted to a standardized T-score using conversion tables published on the PROMIS website (nihpromis.org), with higher scores indicating greater sleep/wake disturbances ^21^.

*ESS:* The ESS score (the sum of 8 item scores, 0-3) ranges from 0-24. The higher the score, the higher that person’s average sleep propensity in daily life or their “daytime sleepiness” ^22^.

8.
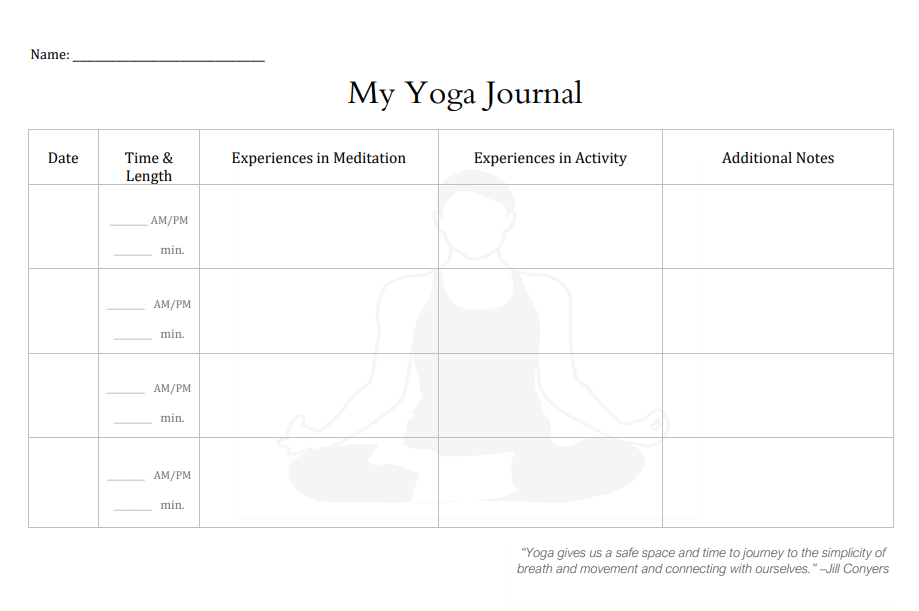
**Journal format for treatment group study participants.**
9. **Yoga session instructor script with pictures**

INTRODUCTION : 5 minutes

**Instructor**: “Welcome to today’s yoga session which should last about an hour. If you need to take a break at any time to use the restroom, cough or just catch your breath, please do so. The yoga practice we will do today is focused on relaxing and stretching and is not designed as vigorous exercise. Our practice will be to incorporate awareness of our breath with very gentle poses to expand our chest and relieve tightness in our muscles.

The session today is divided into 4 parts: 1. A few minutes of settling in, 2. A 25-minute session of gentle stretching postures, 3. A roughly 15-minute period of breathing exercises and 4. A 5-minute guided meditation to end the session.

Feel free to ask questions at any time, to stop the postures or exercises to take a break or just to sit quietly. This is your session.

First off, how are you doing today? Do you feel well enough to participate in the session today? If not, do not worry. You are welcome to stay with the group and just listen and do any of the practice that you feel able to. If you prefer to sign off the call, that is fine too. Again, these sessions are for you.”

SETTLING IN: 5 minutes

**Instructor:** “Take time to get seated comfortably. It’s better is you use a chair with a straight back. If you can, sit forward in the chair in the upright position away from the backrest. If not, it is okay to use the back for support.

It is now time to get in touch with your breath. You are welcome to close your eyes and take off your shoes. Maybe try planting your feet hip width apart and align your ankles directly underneath your knees. Extend your neck with a slight tuck of the chin and lengthen your spine fully. Allow your arms to relax, and rest your palms face up or face down on your thighs. Bring your shoulders back in line with the hips. Keep your chin level and draw it in slightly so the head moves back in line with the shoulders.

We are trying to create a conscious stability from the base of your spine to the crown of your head where you can feel at ease to breathe. Good posture promotes increased breath awareness because there is suddenly much more room to take big, deep breaths.

Keeping your eyes closed, begin to bring your attention to your breath. Just notice where your breath is right now in your body? Be curious about how you are breathing. How long is your inhale? Your exhale? Notice what part of your body you are using most to breathe: your belly, your chest, the back or front of your body, or is it just up top in the neck and shoulders? Let the sounds of the environment fade into the background and let the sound of your breath and the experience of the air moving in and out your lungs come to the foreground. Focus on an open, expansive inhale, filling your lungs from the bottom to the top and a smooth, slow exhale engaging your belly to press the air out of the lungs.

PAUSE to allow time for students to breathe

Allow all the thoughts of your day to fade, bring your attention if it wanders to the breath and the body. Lengthen your inhale and slow down the exhale, breathing out through your mouth. Maintain awareness of your posture, and continue to release excess tension with each exhale, breathe the tension out. If the mind wanders, gently invite your attention back, and follow the experience of the breath moving in and out of the body.

PAUSE again.

And now, gently open your eyes, and prepare for some gentle stretching. If you have any discomfort during these movements, please feel free to ask your instructor to assist you in modifying any of the exercises. If you feel the urge to cough during any of the exercises, feel free to take a rest and do so.

SEATED EXERCISES: 25 minutes

1. SEATED “MOUNTAIN” POSE:

**Instructor:** We will start with the seated mountain pose. Sit with your spine and neck straight and positioned with the shoulders over the hips. Place your hands, palm faced down on your thighs. Take four or five deep breaths in this position.

PAUSE to allow the students to practice.

“Mountain” pose is very restful and is a good point to start each of the other poses from. You can use it as an intermediate stop before you go on to your next pose. It is a very “centering” position for you.

2. SHOULDER CIRCLES:

With your hands at your sides, gently move your shoulders in a circle. Start by making a forward circular motion for 5 repetitions. (INSTRUCTOR DEMONSTRATE)

Then roll the shoulders in the opposite direction for 5 repetitions. (INSTRUCTOR DEMONSTRATE).

Move into your mountain pose for one or two breaths and relax.

3. SUPPORTED NECK STRETCH:

**Instructor:** Next, we will very gently stretch the neck. Place your right hand on the crown of your head with the fingers over the temple. If your shoulder does not feel comfortable in that position, you may leave the arm at your side and stretch the neck gently using the weight of your head. Straighten your left arm down slightly away from your side, keeping the elbow and the wrist extended while stretching out the fingers fully. Gently pull your head to the right and hold for 10 seconds.

PAUSE

Drop your hands to your sides. Repeat the same stretch but this time on the opposite side with your left hand on the crown of your head and your right arm extended to the side. Gently pull your head to the left, while straightening out your right arm slightly away from your side. Keep the elbow and the wrist extended for 10 seconds. Return to mountain pose for two or three relaxing breaths.

4. SEATED “CAT/COW” POSE:

In “cow/cat” pose, move your hands, palms face-down to your knees. Inhale, gently lifting your gaze upward and allowing your spine to arch forward imitating the spine of a cow. Exhale, slowly dropping your chin to your chest, allow your spine to gently round backwards like a cat. This should be a gentle motion, not forced or uncomfortable. It is helpful to synchronize the breath with the movement of the spine. Breath in, gentle arching of the spine while looking up, breath out, gently rounding the spine. Performing 5 to 7 repetitions of arching and rounding, coupled with the breath is ideal.

PAUSE to allow the students to finish.

Bring yourself back to mountain pose and relax.

5. SEATED SIDE BENDS:

**Instructor:** Next we will do seated side bends. Starting from mountain pose, raise your left arm slowly (with your palm facing the ceiling) above your head straight up. Very gently, begin bending laterally to the right. Do not force this. There should be no pinching or discomfort in the spine or ribs. Breathe in and out for 4 or 5 breaths in this position. PAUSE.

You may notice that as you exhale you are able to bend slightly deeper. Gently return to the upright position and bring your left arm down. Now, raise your RIGHT arm up and bend gently to the left side. Again, do not force this bend. Take 4 or 5 breaths in this position.

PAUSE.

Gently return to the upright position and drop your right arm down.

6. SEATED SPINAL TWIST

**Instructor:** “ Our next posture is the seated spinal twist. Starting again from the relaxed mountain pose with your spine straight and long, gently and slowly twist to the left placing your left forearm on the back of your chair. Place your right hand on your left thigh or the left edge of the seat and relax. To deepen the stretch, exhale and GENTLY pull with your right hand. Do not force this. There should be no pain in the spine. If there is any discomfort, back out of this position and tell your instructor. If you are comfortable take 4 or 5 breaths in this position and then return to mountain pose. Begin the twist on the right side by gently twisting to the right and placing your right forearm on the back of your chair. Place your left hand on your right thigh or the edge of the chair and, if you are comfortable, gently deepen the twist by pulling with your left hand. Take 4 or 5 breaths in this position. Return to mountain pose when you are finished. “

Breathing Exercises: 15 minutes

**Instructor:** How do you feel after our gentle yoga poses? If you need to take a drink, relax or change positions, go ahead. In our next exercises, we will be focused on our breath.

1. Counted breaths:

**Instructor:** In this exercise, we will use a metronome that I will set up for us and set a relaxed pace to help you begin to focus on the length of your inhales and exhales. Sitting in a relaxed position with your spine straight (seated mountain pose works great) we will use the metronome to count 3 counts during inhalation and 3 counts during exhalation. Later as we become more experienced, we gradually make the exhalation longer than inhalation but now we will focus on making inhalation and exhalation equal. It is best if you can breathe in and out through your nostrils but if you cannot, through the mouth is fine or in through the nose and out through the mouth.

Instructor: continue this for about one minute and then give the group a break. Any questions?

2. Counted “victory breaths” (ujjayi)

**Instructor:** In this breathing exercise we will be performing the same exercise as we just did but we will be adding something called ujjayi or victory breathing. Victory breathing is performed by very slightly constricting the throat to make a soft sound like waves on the beach. A way to think of the sound is if you are trying to clean a pair of sunglasses by fogging them up before you clean them. DEMONSTRATE TO PARTICIPANTS (i.e. Darth Vader breath or AAAH and HAAA). Take a breath in and exhale like you are trying to fog the glasses and listen to the sound (this is the sound of the victory breath).Then with the same slight constriction of the throat, take a breath in. Again, if possible, you should breath in and out through your nose. Practice taking several breaths in this way and we will then use the metronome and once again count three beats to inhale and three beats to exhale.

Instructor: Have them do this for 45-60 seconds or about 10 breaths. How did that feel? Any questions?

3. Cooling Breath:

Instructor: For this breath technique, which involves drawing air across your tongue and into your mouth, having a “cooling or calming” effect on the nervous system, can be done one of two ways. You can curl the sides of your tongue inward, like you are sucking air through a straw, or if you can’t do that, you can sit your tongue right behind your front teeth and draw air in around your tongue. DEMONSTRATE.

Instructor: Have them practice and once questions are answered have them perform the cooling breath for about 30 seconds.

Brahmari (Buzzing Bee) Breath:

Place your thumbs in your ears and your tips of the fingers over your closed eyelids. Take a deep breath in. Then, while **exhaling, generate a LOUD humming noise so as to feel this vibration resonate through the face and the scalp**. DEMONSTRATE. Make sure you exhale fully allowing the subsequent inhale to be even deeper. Do this for 5 to 7 breaths at your own pace, with the tone that seems most comfortable. Then release the hands onto the thighs and let the breathing settle. Feel the warmth and the calm envelop you.

CLOSING GUIDED MEDITATION: 5 minutes

**Instructor:** Come into a position that is comfortable for you. Let the legs feel heavy. Soften your jaw. Bring your attention once again to your breath. You can breathe slowly keeping inhalation and exhalation slow and in a pattern and rate that feels relaxed and comfortable. Let your whole body melt into the chair, letting go of stiffness or tension in the spine. If your mind becomes distracted by thought, return your awareness to your breath, this inhalation, and this exhalation, being mindful to keep your attention focused in the present moment without trying to change it, and letting everything else be as it is.

Continue to allow your body to relax. Breathe in, breathe out. Take a big cleansing breath and breathe the tension out of your body. You can even make an audible ahhh sound on your next exhale. Let’s try this together, inhale deeply and on your next exhale, open the mouth…ahhhh. One more. Feel the relaxation beginning at the bottom of your feet. Relax your toes.

Let the relaxation spread over your feet and up your ankles, spreading up your calves, and into your knees, softening the backs of your knees and releasing any tension or stiffness in your knees. Feel the relaxation spreading up your legs, feel the tension melt away in your hips and lower back. Release any tension in your lower back. Allow the relaxation to spread upward to your upper back and shoulders. Let go of tension in the shoulders. Feel the relaxation spread up the front of your body starting with your belly, let it be soft, rising and falling with each breath. Let your chest relax, slide your shoulder back, giving you a little more room to breathe easy. Now feel your upper arms relax and let the feeling spread into your elbows and forearms. Let go of tension or stiffness in the hands, relax the palm of your hand, the back of your hand, relax your fingers. Your hands, your arms, feel heavy and warm.

Feel your body unwind even further as you expand your chest open on each inhale and you feel your collar bones widen and relax down. Allow your shoulder blades to ease back slightly and continue to breathe deeply. Deep inhale, slow exhale. Notice your shoulders and your neck, let go of the tension in your neck. Feel the relaxation continue to your chin, let your jaw slacken, release your tongue from the roof of your mouth. Let your eyelids be heavy and relaxed, letting your eyeballs drift back. Notice your eyebrows unfurrowing and relaxing down, let your forehead become smooth and cool; the back of your head relaxes. Let the feeling of relaxation spread to the crown of your head; from the tips of your toes to the top of your head, your entire body is now calm and relaxed. Continue to feel the relaxation and sense of calm flow throughout your body from your head to your feet and back. Notice all of the muscles in your body relaxing completely. Feel your spine lengthen and melt into the chair, your limbs are heavy and warm. Continue to breathe smoothly and slowly as you mentally scan your body, looking for any remaining tension. If you notice any tension, focus on that area and direct the relaxation to flow into that area, carrying away any residual tension.

Relax and return to your Mountain pose. Do you have an questions or thoughts you would like to bring up ? How does everyone feel now that we have completed our session for today ?

**FIGURE LEGENDS**

1. **Figure E1:** Study Protocol.
2. **Figure E2:** Central themes contributing to improved sense of well-being among yoga participants.
3. **Figure E3:** Cough domain score via the Living with Idiopathic Pulmonary Fibrosis scale (L-IPF) (panel A), total score via the Living with Idiopathic Pulmonary Fibrosis scale (L-IPF) (panel B), and cough domain score via the Raghu scale for pulmonary fibrosis (R-Scale-PF) (panel C) from baseline to week 12. Each dot corresponds to a patient, either in the control group (left panel) or the treatment group following the modified yoga program (right panel). Patient observations are overlaid on top of violin plots and displayed twice within each panel: once at baseline and again at week 12. Lower scores represent lesser symptoms/impairment.

**TABLES**

**Table E1:** Description of yoga poses.

| **Pose name** | **Description** | **Frequency** | **Duration** |
| --- | --- | --- | --- |
| **Asanas (postures)** | | | |
| Parvatasana (seated mountain pose) | Straight back, lengthen up spine, relax shoulders down, neutral chest/diaphragm, deep breaths | Starting point prior to subsequent poses | 2 min |
| Chair seated shoulder circles | Roll shoulders in a circular motion (both directions, 5 repetitions) | 1 round | 2 min |
| Seated neck stretches | Gently pull head to each side and hold | 1 round | 2 min |
| Marjayasana-bitilasana (seated cat cow pose) | Inhale and arch spine forward, exhale and round spine backwards | 5-7 repetitions | 4 min |
| Parsva Sukhasana (seated side bend pose) | Raise arm and bend laterally to each side | 1 round | 2 min |
| Ardha Matsyendrasana (seated spinal twist) | Twist to each side, rotating the spine, opening shoulders and chest | 1 round | 3 min |
| **Pranayama (breathing exercises)** | | | |
| Counted breaths | Through nostrils, 3 counts inhalation, 3 counts exhalation using metronome to set pace | 1 round | 1 min |
| Ujjayi (counted “victory breaths” | As above, adding throat constriction | 1 round | 1 min |
| Cooling breath | Curl sides of tongue inward, inhale, exhale | 1 round | 30 sec |
| Brahmari (buzzing bee breath) | Place thumbs in ears and fingers over closed eyelids, inhale, exhale with loud humming noise | 1 round | 1 min |
| **Dhyana (meditation)** | | | |
| Guided mediation | N/A | 1 round | 5 min |

**Table E2:** Changes from baseline to week 12 and adjusted treatment effect of modified yoga program on secondary endpoints for the study population.

|  | *Baseline*  [*] | *Week 12*  [*] | *Change* [†]  *(Week 12 – Baseline)* | *Adjusted Treatment Effect of Yoga* [‡] | |
| --- | --- | --- | --- | --- | --- |
|  |  |  |  | *Point estimate*  *(95% CI)* | *p-value* |
| **Absolute Forced Vital Capacity (FVC) (L)** | | | | | |
| MYP | 2.95 (0.86) | 2.82 (0.86) | -0.11 (0.23) | 0.02  (-0.19, 0.23) | 0.88 |
| Control | 2.67 (0.77) | 2.54 (1.02) | -0.13 (0.54) |  |  |
| **Percent Predicted Forced Vital Capacity (FVC) (%)** | | | | | |
| MYP | 79.57 (20.28) | 77.07 (21.97) | -2.55 (6.09) | 0.25  (-6.99, 7.49) | 0.94 |
| Control | 70.03 (21.84) | 70.63 (24.80) | 0.60 (19.06) |  |  |
| **Absolute Diffusing capacity or transfer factor of the lung for carbon monoxide (DLCO) (ml/min/kPa)** | | | | | |
| MYP | 13.27 (4.42) | 12.08 (3.93) | -1.18 (1.88) | -0.41  (-1.66, 0.84) | 0.51 |
| Control | 12.38 (3.41) | 11.50 (4.60) | -0.85 (2.66) |  |  |
| **Percent Predicted Diffusing capacity or transfer factor of the lung for carbon monoxide (DLCO) (%)** | | | | | |
| MYP | 56.03 (20.97) | 52.48 (20.28) | -3.11 (12.03) | -0.73  (-7.10, 5.63) | 0.82 |
| Control | 52.47 (15.38) | 49.79 (19.98) | -2.28 (11.25) |  |  |
| **6-minute walking test (6MWT) (feet)** | | | | | |
| MYP | 1268.79 (403.62) | 1195.37 (383.21) | -69.92 (143.71) | 83.49  (-30.09, 197.08) | 0.15 |
| Control | 1311.07 (405.74) | 1166.76 (526.65) | -146.07 (279.52) |  |  |
| **Notes:**  [*] Week 12 is the end of the intervention period. Mean (standard deviation) values were computed for all patients with complete data at baseline and week 12.  [†] We perform an unadjusted comparison of the change (week 12 – baseline) of each of the primary outcomes between the two study arms (treatment and control).  [‡] A linear regression model was used treatment differences at week 12 for outcomes, adjusting for age, sex, BMI, supplementary oxygen usage, and the baseline value of the specific outcome measure under consideration. To handle missing data, multiple imputation was performed using the chained equations method. Pooled estimates, confidence intervals (CIs), and p-values were obtained using Rubin’s rules (N=60). | | | | | |

**Table E3: Complete case analysis for changes from baseline to week 12 and adjusted treatment effect of modified yoga program on primary endpoints for the study population.**

|  | *Baseline** | *Week 12** | *Change*†  *(Week 12 – Baseline)* | *Adjusted Treatment Effect of Yoga*‡ | |
| --- | --- | --- | --- | --- | --- |
|  |  |  |  | *Point estimate*  *(95% CI)* | *p-value* |
| **Living with Idiopathic Pulmonary Fibrosis (L-IPF) (Cough)** | | | | | |
| MYP | 27.83 (25.69) | 18.10 (21.23) | -9.31 (21.03) | -8.59  (-17.50, 0.33) | 0.065 |
| Control | 24.83 (22.95) | 25.00 (22.09) | 0.17 (19.14) |  |  |
| **Living with Idiopathic Pulmonary Fibrosis (L-IPF) (Dyspnea)** | | | | | |
| MYP | 24.14 (21.81) | 21.27 (20.02) | -1.61 (12.09) | -2.93  (-8.77, 2.90) | 0.329 |
| Control | 21.27 (20.02) | 22.86 (22.48) | 0.99 (11.03) |  |  |
| **Living with Idiopathic Pulmonary Fibrosis (L-IPF) (Overall)** | | | | | |
| MYP | 30.84 (18.77) | 23.37 (16.49) | -7.18 (13.57) | -7.14  (-13.06, -1.23) | 0.022 |
| Control | 26.49 (19.60) | 27.16 (19.92) | 0.66 (10.70) |  |  |
| **Raghu Scale for Pulmonary Fibrosis (R-scale-PF) (Cough)** | | | | | |
| MYP | 3.20 (2s.64) | 2.07 (2.31) | -0.97 (2.29) | -1.19  (-2.24, -0.14) | 0.031 |
| Control | 2.87 (2.21) | 3.14 (2.56) | 0.28 (2.05) |  |  |
| **Raghu Scale for Pulmonary Fibrosis (R-scale-PF) (Shortness of Breath)** | | | | | |
| MYP | 4.13 (2.62) | 4.00 (2.58) | 0.00 (1.71) | -0.08  (-1.01, 0.84) | 0.859 |
| Control | 3.27 (2.48) | 3.66 (2.96) | 0.38 (1.90) |  |  |
| **Raghu Scale for Pulmonary Fibrosis (R-scale-PF) (Fatigue)** | | | | | |
| MYP | 4.43 (2.45) | 4.31 (2.88) | 0.00 (2.20) | 0.09  (-1.05, 1.22) | 0.884 |
| Control | 3.67 (2.66) | 3.83 (3.08) | 0.14 (2.08) |  |  |
| **Raghu Scale for Pulmonary Fibrosis (R-scale-PF) (Mood)** | | | | | |
| MYP | 2.63 (2.61) | 1.90 (2.54) | -0.79 (2.46) | -0.18  (-1.30, 0.95) | 0.757 |
| Control | 1.63 (1.92) | 1.48 (2.11) | -0.07 (2.09) |  |  |
| **Raghu Scale for Pulmonary Fibrosis (R-scale-PF) (Wellbeing)** | | | | | |
| MYP | 3.03 (1.88) | 3.21 (2.35) | 0.07 (2.46) | -0.54  (-1.79, 0.70) | 0.395 |
| Control | 2.80 (2.04) | 3.48 (2.95) | 0.72 (2.17) |  |  |
| **Raghu Scale for Pulmonary Fibrosis (R-scale-PF) (Overall)** | | | | | |
| MYP | 17.43 (9.63) | 15.48 (9.88) | -1.69 (8.65) | -2.12  (-6.39, 2.16) | 0.336 |
| Control | 14.23 (8.84) | 15.59 (10.93) | 1.45 (7.60) |  |  |
| **King’s Brief Interstitial Lung Disease (K-BILD) (Psychological)** | | | | | |
| MYP | 36.40 (10.52) | 38.45 (9.10) | 1.90 (8.51) | 0.33  (-3.85, 4.50) | 0.879 |
| Control | 40.20 (10.04) | 40.17 (10.18) | -0.03 (9.95) |  |  |
| **King’s Brief Interstitial Lung Disease (K-BILD) (Breathlessness and Activities)** | | | | | |
| MYP | 24.03 (8.72) | 24.21 (7.89) | 0.14 (4.76) | 0.05  (-2.09, 2.20) | 0.960 |
| Control | 25.57 (8.59) | 25.63 (7.89) | 0.07 (4.62) |  |  |
| **King’s Brief Interstitial Lung Disease (K-BILD) (Chest Symptoms)** | | | | | |
| MYP | 11.27 (2.21) | 11.93 (2.2) | 0.62 (1.90) | 0.27  (-0.54, 1.08) | 0.514 |
| Control | 11.70 (2.31) | 12.00 (2.12) | 0.30 (1.58) |  |  |
| **King’s Brief Interstitial Lung Disease (K-BILD) (Overall)** | | | | | |
| Yoga | 71.73 (17.79) | 74.59 (17.27) | 2.62 (11.73) | 1.09  (-5.11, 7.28) | 0.733 |
| Control | 77.57 (18.33) | 77.80 (17.64) | 0.23 (13.94) |  |  |
| **EuroQol-5D-5L (EQ-5D-5L) Visual Analogue Scale (VAS)** | | | | | |
| MYP | 70.53 (18.77) | 70.76 (16.71) | 0.90 (13.27) | 2.62  (-3.44, 8.68) | 0.401 |
| Control | 71.57 (21.88) | 68.43 (21.66) | -3.13 (12.40) |  |  |
| **EuroQol-5D-5L (EQ-5D-5L) Index** | | | | | |
| MYP | 0.81 (0.11) | 0.80 (0.16) | -0.01 (0.11) | -0.00  (-0.05, 0.04) | 0.837 |
| Control | 0.82 (0.10) | 0.82 (0.12) | 0.00 (0.07) |  |  |
| **Hospital Anxiety and Depression Scale (HADS) (Anxiety)** | | | | | |
| MYP | 4.80 (4.13) | 3.90 (3.72) | -0.90 (2.87) | -0.10  (-1.22, 1.02) | 0.860 |
| Control | 3.23 (2.49) | 2.93 (2.66) | -0.17 (2.19) |  |  |
| **Hospital Anxiety and Depression Scale (HADS) (Depression)** | | | | | |
| MYP | 4.57 (3.82) | 3.76 (3.52) | -0.90 (3.07) | -0.84  (-2.17, 0.48) | 0.218 |
| Control | 4.17 (2.90) | 4.28 (3.5) | 0.17 (2.16) |  |  |
| **Hospital Anxiety and Depression Scale (HADS) (Overall)** | | | | | |
| MYP | 9.37 (7.45) | 7.66 (6.68) | -1.79 (4.97) | -1.04  (-3.16, 1.09) | 0.345 |
| Control | 7.40 (4.80) | 7.17 (5.51) | -0.03 (3.59) |  |  |
| **Patient-Reported Outcomes Measurement Information System (PROMIS™) Sleep Disturbance (SD)** | | | | | |
| MYP | 45.29 (9.52) | 45.80 (8.73) | 0.89 (8.60) | 2.35  (-1.10, 5.79) | 0.188 |
| Control | 48.62 (9.48) | 46.29 (9.6) | -2.13 (5.76) |  |  |
| **Patient-Reported Outcomes Measurement Information System (PROMIS™) Sleep-Related Impairment (SRI)** | | | | | |
| MYP | 47.32 (8.78) | 45.77 (8.08) | -1.10 (7.62) | -1.77  (-5.50, 1.95) | 0.356 |
| Control | 44.63 (9.61) | 45.92 (10.66) | 1.58 (7.66) |  |  |
| **Epworth Sleepiness Scale (ESS)** | | | | | |
| MYP | 7.03 (3.94) | 6.03 (4.40) | -0.90 (2.92) | -0.79  (-2.49, 0.91) | 0.367 |
| Control | 4.63 (2.92) | 4.90 (3.57) | 0.38 (3.16) |  |  |
| **Notes:**  [*] Week 12 is the end of the intervention period. Mean (standard deviation) values were computed for all patients with complete data at baseline and week 12.  [†] We perform an unadjusted comparison of the change (week 12 – baseline) of each of the primary outcomes between the two study arms (treatment and control).  [‡] A linear regression model was used treatment differences at week 12 for outcomes, adjusting for age, sex, BMI, supplementary oxygen usage, and the baseline value of the specific outcome measure under consideration. Complete case analysis results presented here. | | | | | |

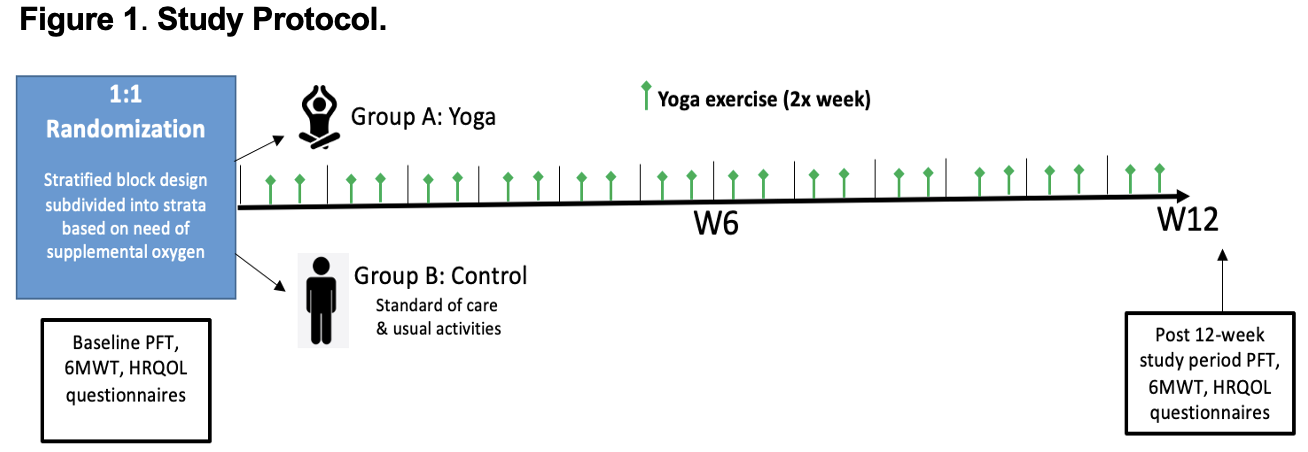
**FIGURES**

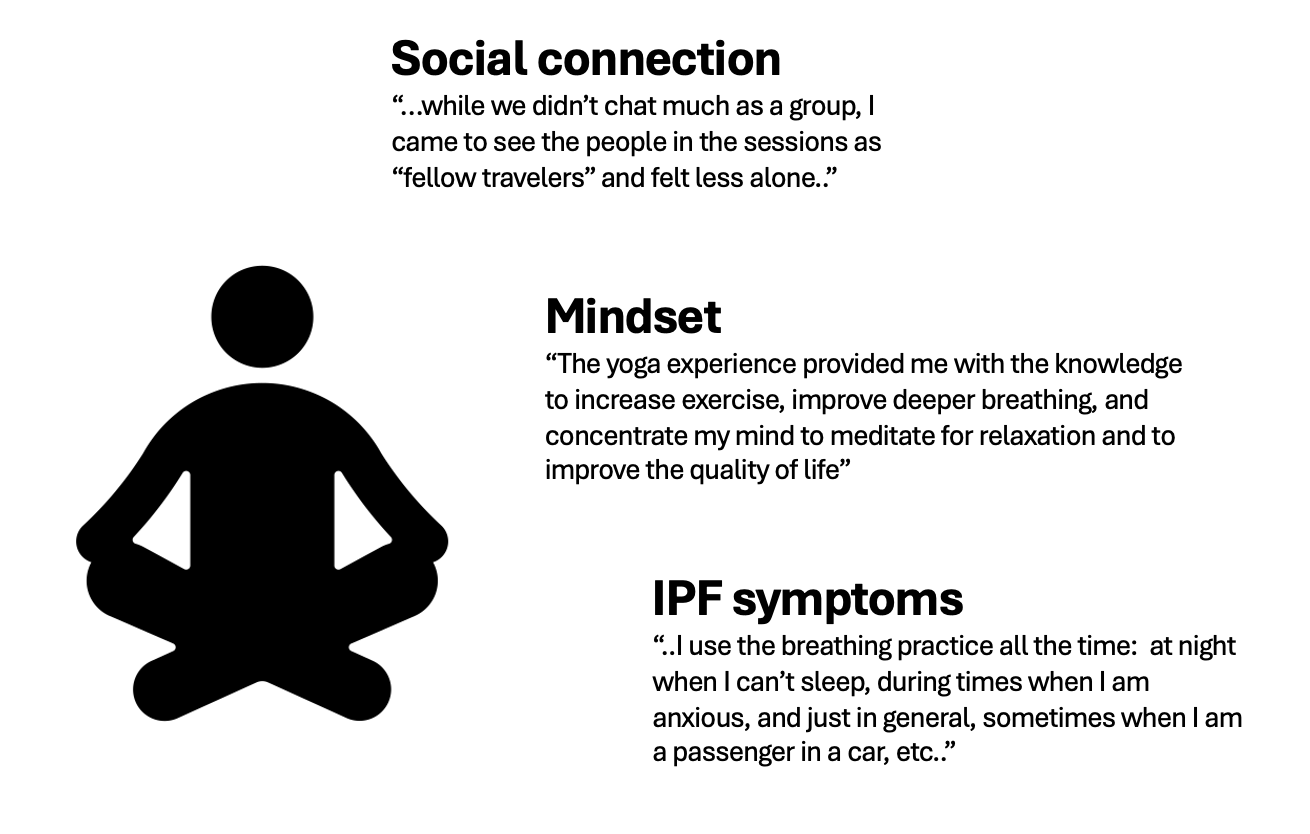
**Figure E1:** Study protocol.

**Figure E2:** Central themes contributing to improved sense of well-being among yoga participants.

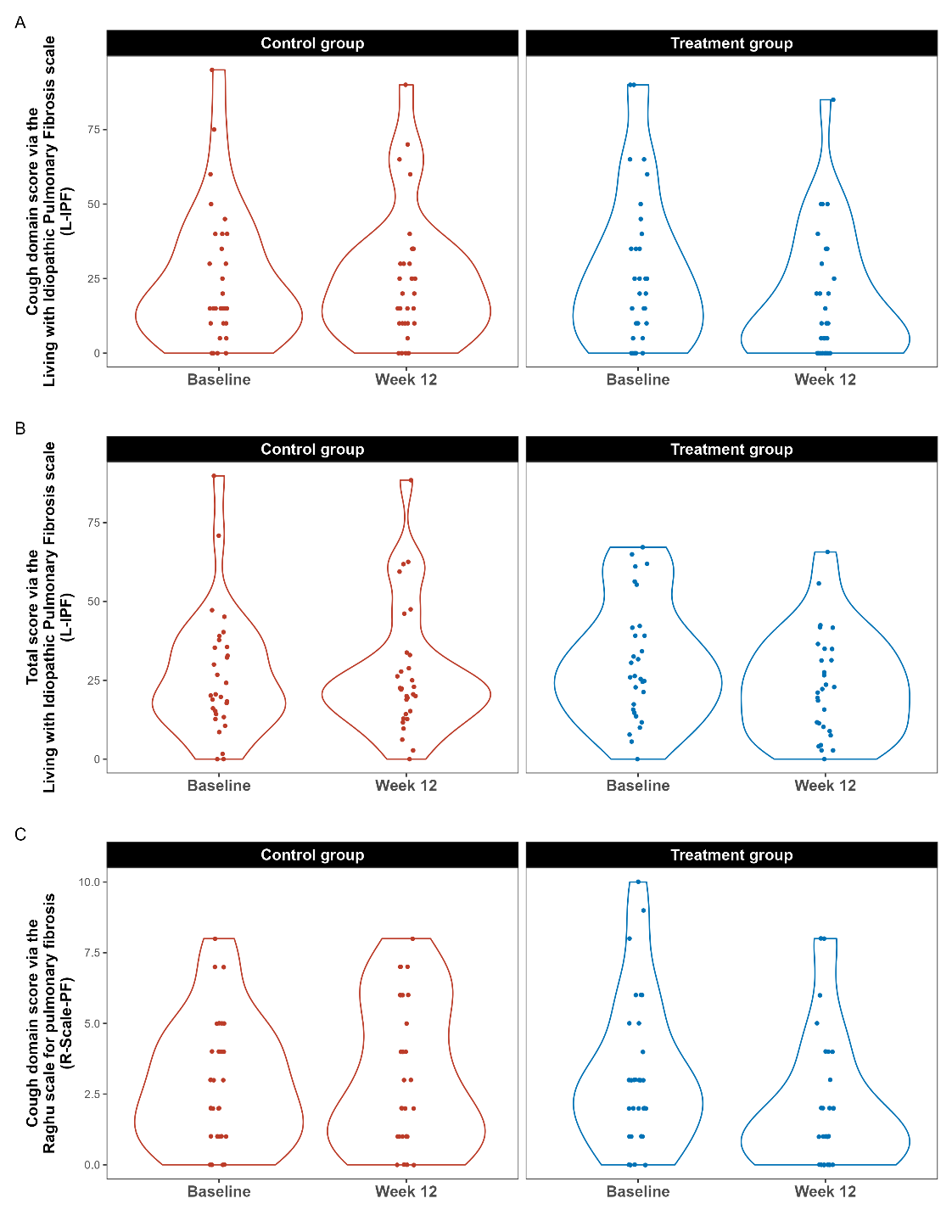
**Figure E3.** Cough domain score via the Living with Idiopathic Pulmonary Fibrosis scale (L-IPF) (panel A), total score via the Living with Idiopathic Pulmonary Fibrosis scale (L-IPF) (panel B), and cough domain score via the Raghu scale for pulmonary fibrosis (R-Scale-PF) (panel C) from baseline to week 12.
